# Development Efficiency and Mortality After Coronary Artery Bypass Grafting: A National Causal Inference Analysis

**DOI:** 10.64898/2026.02.02.26345421

**Authors:** Gabriel K. Martins, Arthur D. Botelho, Leo Consoli, Iago T. Grillo, Felipe S. Passos, Ricardo E. Treml, Tulio Caldonazo

## Abstract

**Background:** We evaluated whether development efficiency, the component of the Human Development Index independent of GDP per capita and income inequality, is associated with in-hospital mortality after coronary artery bypass grafting (CABG) in Brazil, and whether this association is mediated by access to elective surgery.

**Methods:** We conducted a retrospective ecological panel study using administrative data on CABG hospitalizations within the Brazilian Unified Health System from 2008-2024. State-year observations were linked to socioeconomic indicators. Development efficiency was defined as the residual of HDI after regression on GDP per capita and the Gini coefficient. Associations with in-hospital mortality were examined using volume-weighted multilevel models. Absolute causal effects and mediation through urgency status were estimated using g-computation and parametric causal mediation analysis.

**Results:** The final analytic panel included 379 state-year observations. A 1–standard deviation increase in development efficiency was associated with a reduction in predicted in-hospital mortality from 6.8% to 5.7% (absolute risk reduction −1.1 percentage points; p<0.001), corresponding to one death prevented for every 91 procedures. Mediation analysis indicated that 95.5% of the total effect was attributable to the natural direct effect, while only 4.5% was mediated through urgency status, with no significant indirect effect.

**Conclusions:** Development efficiency is an independent and clinically meaningful determinant of survival after CABG in Brazil. Higher income-independent HDI performance is associated with substantial absolute mortality reductions, driven predominantly by direct system-level pathways rather than changes in urgency profile. Strengthening health-system efficiency and perioperative capacity may therefore yield meaningful gains in cardiac surgical outcomes.

## INTRODUCTION

Coronary artery disease remains the leading cause of death worldwide, accounting for approximately 17.5 million deaths each year, with nearly 80% occurring in low- and middle-income countries.[1] Coronary artery bypass grafting (CABG) is the gold-standard treatment for complex multivessel disease and provides durable survival benefits in high-risk populations.[2-7] Despite its effectiveness, global access to cardiac surgery remains highly unequal. An estimated 75% of the world’s population lacks timely access to CABG when clinically indicated, largely due to constraints in infrastructure, workforce capacity, and health-system organization rather than clinical contraindications. [1,4,6] Marked differences in specialist availability further illustrate this imbalance, with low-income countries averaging approximately 0.04 cardiac surgeons per million inhabitants versus more than 7 per million in high-income settings.[1,8] Consistently, population-level analyses show that surgical mortality is inversely associated with the Human Development Index (HDI), indicating that outcomes after cardiac surgery are shaped not only by patient risk but also by broader social and institutional conditions.[1,4,7,13]

However, important uncertainties remain regarding how development influences surgical survival. Most prior investigations of disparities in cardiac surgery outcomes have focused on macroeconomic indicators such as gross domestic product per capita or income inequality (the Gini coefficient).[9] While useful, these measures do not capture how effectively societies convert available resources into health, education, and longevity.[8,9] Notably, nearly one-third of the variation in HDI is independent of income and inequality, representing a dimension of development efficiency that may more directly reflect health-system performance. [1,9] The relationship between this income-independent component of development and CABG mortality has not been formally evaluated using causal inference methods, and the mechanisms linking development to perioperative outcomes remain insufficiently characterized.

Brazil provides a uniquely informative setting to examine these relationships. The Brazilian Unified Health System (Sistema Único de Saúde, SUS) is one of the largest universal public health systems worldwide and offers nationwide coverage for cardiac surgery. Despite universal entitlement, the delivery of CABG varies by more than 700-fold across states, reflecting deep heterogeneity in surgical capacity, workforce distribution, and perioperative infrastructure. [8,14] This combination of universal coverage with large regional variation creates a natural analytic framework to evaluate how development-related factors translate into differences in surgical outcomes within a single national system. In this study, we tested the hypothesis that the income-independent component of development efficiency causally reduces in-hospital mortality after CABG in Brazil, and that its protective effect is driven predominantly by direct health-system mechanisms rather than solely by improved access to elective surgery. [1,9]

## METHODS

### Data Sources and Study Design

We performed a retrospective, population-based ecological panel study using administrative data on CABG hospitalizations within the Brazilian Unified Health System (SUS) from January 1, 2008, through December 31, 2024. Surgical data were obtained from the SUS Hospital Information System (SIH/SUS), which captures all publicly funded inpatient procedures nationwide.

### Study Population and Variables

CABG hospitalizations were identified using national procedure codes for coronary revascularization (04.06.01.092-7, 04.06.01.093-5, 04.06.01.094-3, 04.06.01.095-1). The outcomes included were in-hospital mortality and procedures volume.

Key covariates included revascularization extent, urgency status (proportion of urgent/emergency procedures), calendar year, and geographic macroregion (North, Northeast, Southeast, South, Central-West), modeled as a hierarchical level to account for regional clustering and health-system heterogeneity.

Hospitalization records were aggregated at the state-year level (27 states over multiple years) and stratified by procedure type and urgency status. This aggregation strategy allowed us to capture temporal and geographic variation in surgical volume and outcomes while preserving comparability across states within the public health system.

### Socioeconomic Exposures

Socioeconomic indicators were derived from national public databases. HDI values were obtained from the Brazilian Institute of Geography and Statistics for the years 2012-2021 and interpolated within states for earlier years (2000-2011) using linear interpolation. Annual illiteracy rates (2012-2021); gross domestic product (GDP) per capita (2013-2016); and the Gini coefficient (2000 and 2010) were similarly interpolated to generate complete annual state-level, as the complete availability for public data for those years were not possible from official sources. To ensure temporal ordering, all socioeconomic exposures were lagged by one year relative to surgical outcomes, establishing temporal precedence and minimizing the potential for reverse causation.

### Development Efficiency Decomposition

To separate development performance from pure economic resources, lagged HDI was regressed on lagged GDP per capita and lagged Gini coefficient. The residual from this model, accounting for 28.6% of HDI variance (adjusted R^2^ = 0.7137), was defined as development efficiency, representing the capacity of social and institutional systems to convert available resources into health, education, and longevity, independent of financial inputs.

### Statistical Analysis

To account for heteroskedasticity and large inter-state variability in surgical volume (>700-fold), state-year observations were volume-weighted to stabilize mortality estimates. Multilevel regression models were fitted with state-year as the analytic unit and macroregion as a higher-level grouping factor. Weights were defined as the number of CABG procedures in each state-year relative to the total study volume, so that states with higher-volume programs, with more stable mortality estimates, contributed proportionally more to model fitting and small-volume states had reduced influence from stochastic variation.

Absolute causal effects were estimated using g-computation with covariates held at observed values. Causal mediation analysis was conducted using quasi-Bayesian simulation (1,000 draws) to decompose total effects into natural direct and indirect components mediated by urgency proportion.

Sensitivity analyses included: (1) restriction to high-volume states (mean procedures ≥75th percentile); (2) elective-only procedures; (3) non-interpolated data only; and (4) alternative time periods. Spatial autocorrelation was assessed using Moran’s I. Robustness to unmeasured confounding was evaluated with E-values. Multicollinearity was checked via variance inflation factors (VIF), ensuring it was <5 in all models.

All analyses were performed in R. Two-sided P <0.05 was considered statistically significant, with emphasis on absolute effects and clinical interpretability given the rarity of mortality [1]. Absolute effects were calculated to enhance clinical interpretability. The Absolute Risk Reduction (ARR) was estimated using g-computation by calculating the difference between the marginal predicted mortality at the baseline HDI residual and the predicted mortality following a one-standard deviation (1-SD) increase in development efficiency. The Number Needed to Treat (NNT), representing the number of CABG procedures required in a high-efficiency environment to prevent one additional in-hospital death compared to a low-efficiency one, was calculated as the inverse of the absolute risk reduction (NNT=1/ARR).

### Handling of Missing Data and Interpolation

Missing socioeconomic data was addressed via linear interpolation within states, ensuring a complete panel. Interpolation flags were tracked, and sensitivity analyses restricted to non-interpolated years showed consistent results.

### Ethical Considerations

The study used only de-identified, aggregated public data from the SIH/SUS database. No individual-level identifiable information was accessed, and all analyses were conducted at the state-year level without direct involvement of human participants. In accordance with Brazilian regulations, studies based exclusively on anonymized secondary data from publicly available databases are exempt from Research Ethics Committee review, as established by National Health Council Resolution No. 510/2016.

## RESULTS

### Study Population

The final analytic dataset included 379 state-year observations of CABG hospitalizations in the SUS from 2008 to 2024, representing more than 96% of eligible CABG records after lagging and completeness filters (excluding state-years without procedures). Substantial heterogeneity was observed across states in both socioeconomic development and CABG outcomes.

### Absolute Causal Effect

Using g-computation with parametric simulation–based causal mediation, a 1–standard deviation increase in development efficiency was associated with a reduction in predicted in-hospital mortality from 6.8% to 5.7%, corresponding to an absolute risk reduction of −1.1 percentage points (95% CI −1.53 to −0.61; p < 0.001; Table 1). Effect decomposition indicated that the natural direct effect accounted for most of the survival benefit, while the proportion of urgent procedures did not significantly mediate the association (Figure 1 and Table 2).

**Table 1.** Absolute Causal Effect of a 1-Standard Deviation Increase in HDI Residual on In-Hospital CABG Mortality (G-Computation)

| Quantity | Estimate |
| --- | --- |
| Baseline mortality (low HDI residual) | 6.8% |
| Mortality after + 1-SD in HDI residual | 5.7% |
| Absolute risk reduction | -1.1% (95% CI: -1.53% to -0.61%) |
| P value | <0.001 |
| Number needed to treat (procedures per life saved) | 91 |

**Footnote:** Estimates from g-computation holding constant surgical risk factors, urgency mix, procedure type, calendar year, and regional structure. Absolute risk reduction interpreted as percentage points.

**Table 2.** Decomposition of the Effect of HDI Residual on CABG Mortality via Proportion of Urgent Procedures.

| Effect | Risk Difference | 95% Confidence Interval | P Value |
| --- | --- | --- | --- |
| Total effect | −1.10% | −1.53% to −0.61% | <0.001 |
| Natural direct effect (NDE/ADE) | −1.02% | −1.47% to −0.58% | <0.001 |
| Natural indirect effect (NIE/ACME) | −0.05% | −0.16% to 0.01% | 0.108 |
| Proportion mediated (average) | 4.5% | −7% to 15% | 0.108 |

**Footnote:** NDE = natural direct effect; NIE = natural indirect effect; ACME = average causal mediation effect. Mediation via proportion of urgent CABG procedures. Estimates from parametric simulation-based approach (quasi-Bayesian).

**Figure 1.**
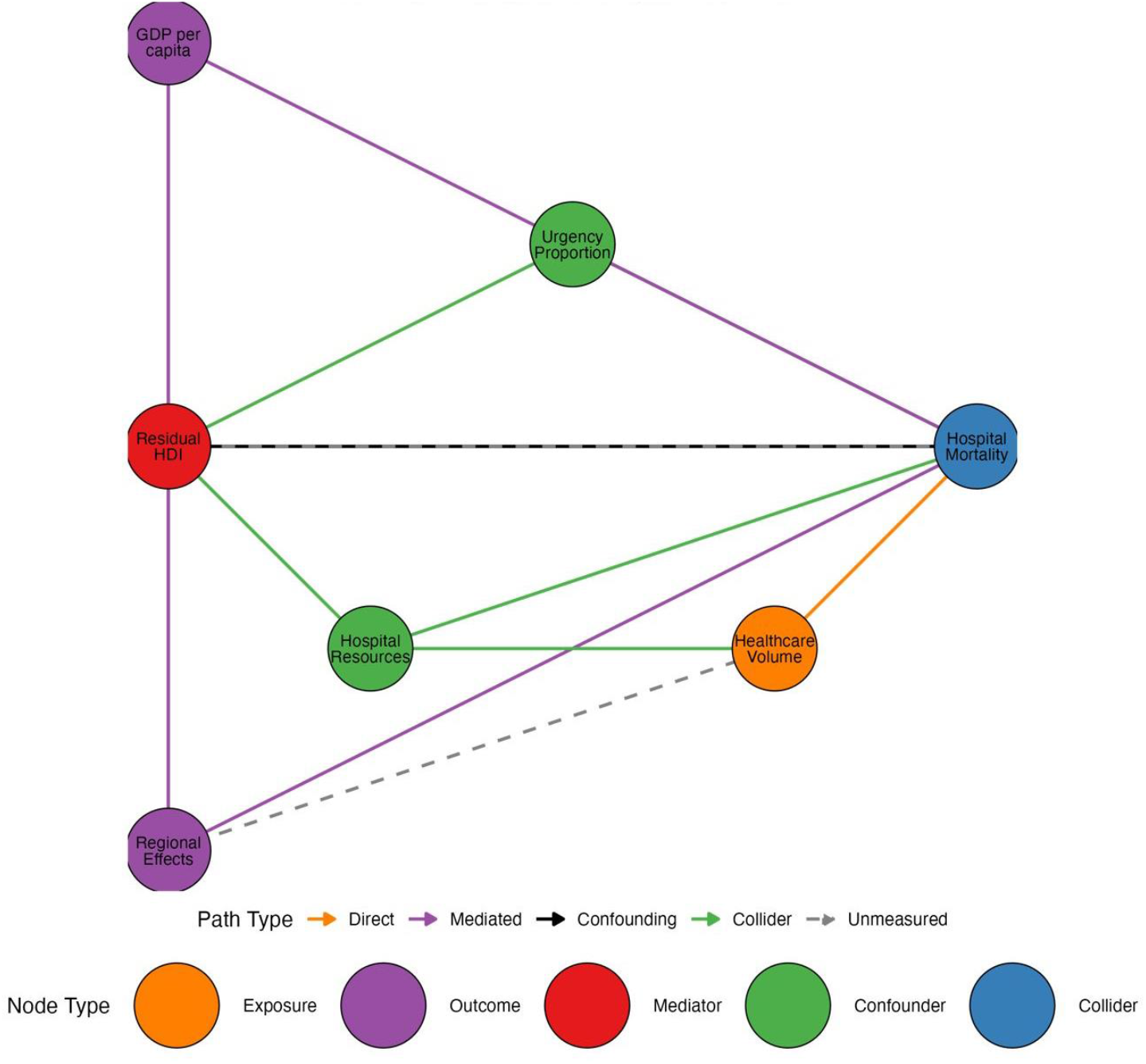
Directed Acyclic Graph of the Causal Pathways Between Development Efficiency (Residual HDI) and In-hospital Mortality Following Coronary Artery Bypass Grafting This causal diagram maps the hypothesized relationships between socioeconomic exposures, system-level mediators, and surgical outcomes. Development Efficiency (Residual HDI), defined as the component of the Human Development Index independent of income and inequality, serves as the primary exposure. Urgency Proportion and Hospital Resources are identified as potential mediators through which development may influence survival. GDP per capita and Regional Effects are treated as confounding variables to account for economic resources and geographic health-system heterogeneity.

### Mediation by Surgical Urgency

Mediation analysis showed that the natural direct effect accounted for nearly all the observed benefit (−1.02%, 95% CI −1.47% to −0.58%; p < 0.001), whereas the natural indirect effect through urgency proportion was small and not statistically significant (−0.05%, 95% CI −0.16% to 0.01%; p = 0.108). The proportion mediated was 4.5% (95% CI −7% to 15%), indicating that most of the total effect was direct rather than mediated by urgency status.

### Clinical and Population-Level Implications

At the national level, given approximately 40,000 annual CABG within SUS, a 1-SD increase in development efficiency would be expected to prevent roughly 440 in-hospital deaths per year (Figure 2).

**Figure 2.**
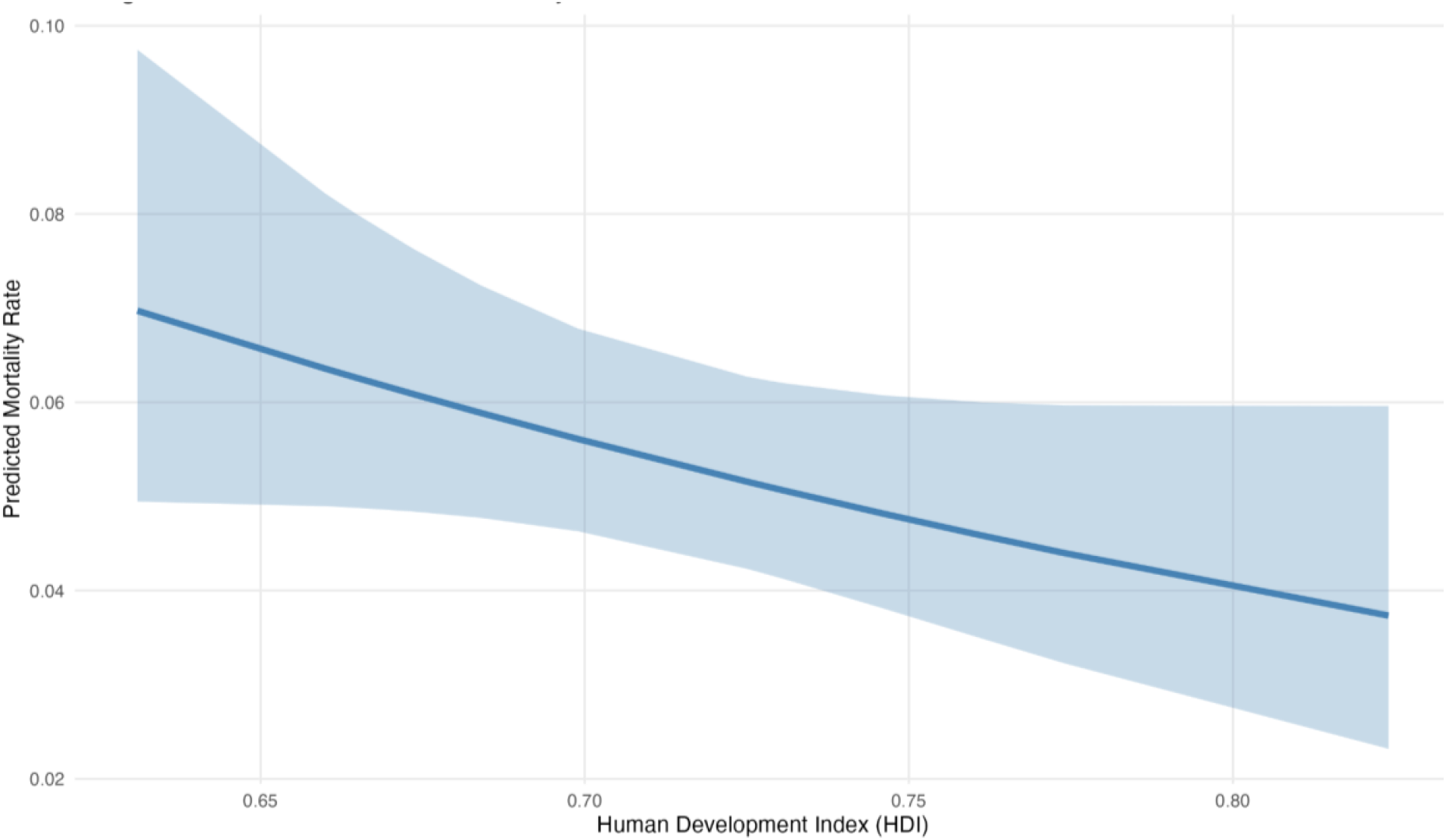
Marginal Effect of the Human Development Index on Predicted CABG Mortality Rate This figure illustrates the predicted probability of in-hospital mortality following coronary artery bypass grafting (CABG) as a function of the Human Development Index (HDI). The solid blue line represents the marginal predicted mortality rate, showing a clear inverse relationship where higher development levels are associated with lower mortality. The shaded area indicates the 95% confidence interval. According to the predictive model, a 1-standard deviation increase in development efficiency leads to a significant absolute reduction in mortality from 6.8% to 5.7%.

### Robustness of the Causal Association

The association remained stable across sensitivity analyses, including restriction to high-volume centers, elective procedures only, and alternative time-period specifications. No meaningful spatial autocorrelation was detected (Moran’s I = −0.002; p = 0.31). The E-value was 1.66 for the point estimate and 2.26 for the lower confidence bound, indicating that an unmeasured confounder would need to be more strongly associated with both development efficiency and mortality than most known clinical risk factors to fully explain the observed effect.

## DISCUSSION

In this nationwide longitudinal analysis of CABG within the Brazilian SUS, higher development efficiency, measured as the income-independent component of the HDI, was associated with lower in-hospital mortality. Each 1-SD increase in this measure corresponded to an absolute 1.1% decrease in mortality, equivalent to one life saved for every 91 operations and approximately 440 deaths being prevented annually at the national level. This effect size is clinically meaningful and comparable to that of major therapeutic advances in cardiovascular care.[10]

More than 95% of the observed survival benefit was not mediated by reductions in urgent surgery. Although urgent CABG is associated with substantially higher mortality,[11,12] the effect of development efficiency appears to operate predominantly through direct system-level mechanisms. This interpretation is supported by evidence that institutional experience, perioperative organization, ICU staffing, and structured quality programs are major determinants of cardiac surgical outcomes.[14–17] High-volume centers show 13–15% lower mortality than low-volume hospitals,[18] and intensivist-led cardiac ICUs and formal quality-improvement programs reduce postoperative mortality by 30–50%, independent of surgical timing.[19–21] Together, these data indicate that survival depends more on perioperative system capacity and organization than on urgency status alone.

The HDI residual reflects dimensions such as education, institutional capacity, and health-system functionality that are not captured by income alone. Low health literacy, limited workforce density, and inadequate hospital preparedness are each independently associated with worse cardiovascular and surgical outcomes.[22–28] Regions with fewer than 20–40 surgical specialists per 100,000 inhabitants show substantially poorer surgical and obstetric results, and global estimates indicate that workforce expansion could avert hundreds of thousands of deaths annually.[24–26] At the hospital level, reliable infrastructure, medication availability, organized referral pathways, dedicated teams, and perioperative quality programs are consistently associated with lower mortality and failure-to-rescue rates, with reported reductions of up to 50%.[20,27–33] Together, these system-capacity domains closely align with what is captured by the HDI residual.

Taken together, these findings carry important policy implications. In a middle-income country with universal health coverage, development efficiency functions as a structural determinant of cardiac surgical survival. Gains in this domain have the potential to save hundreds of lives annually after CABG by strengthening the system-level pathways identified in our mediation analysis. Evidence-based strategies include regionalization to high-volume centers,[13,14,34,35] minimum surgical workforce thresholds,[24,25,35] structured perioperative quality programs such as ERAS and care bundles,[31,32,35] health-literacy interventions to support recovery,[22,33] and expansion of dedicated, intensivist-led cardiac intensive care units.[19,20,34] Investing in these organizational and capacity-building measures offers a scalable route to improving surgical outcomes and links social and institutional development directly to cardiovascular survival.

## Study Limitations

This study has several important limitations. First, the ecological design introduces a risk of ecological fallacy, as associations observed at the state-year level may not fully reflect individual-level relationships. Second, the use of administrative data limits clinical granularity, precluding adjustment for patient-level risk factors such as age, comorbidities, coronary anatomy, and operative complexity, and preventing center-level analyses of hospital performance. Third, socioeconomic series required interpolation for several variables, particularly GDP per capita and the Gini coefficient, which may introduce measurement error and temporal smoothing bias. Fourth, changes in SIH/SUS coding practices over time may have affected procedure classification and outcome reporting. Fifth, administrative data do not reliably distinguish isolated CABG from combined procedures, which may contribute to residual confounding. Finally, differential reporting quality across states is possible, such that more developed systems may capture outcomes more completely, introducing potential information bias and endogeneity. Despite these constraints, the consistency of results across multiple sensitivity analyses and their concordance with the broader surgical outcomes literature support the robustness of the main findings.

## CONCLUSION

Development efficiency is an independent and clinically important determinant of survival after CABG in Brazil. Improvements in the income-independent component of HDI are associated with substantial absolute reductions in in-hospital mortality, largely through direct system-level pathways rather than shifts in urgency status. Investments in health-system efficiency and perioperative capacity are therefore central to improving cardiac surgical outcomes.

## Data Availability

All the data is available publicly

## Acknowledgements

None

